# “If all of this was about HEALTH, I’d still be working”: Lived Experiences of Covid-19 Vaccine Mandates of Healthcare Workers in British Columbia, Canada

**DOI:** 10.1101/2025.05.22.25328161

**Authors:** Claudia Chaufan, Natalie Hemsing, Rachael Moncrieffe

## Abstract

**Background:** COVID-19 vaccine mandates for healthcare workers (HCWs) in the Canadian province of British Columbia (BC) were implemented in October 2021. Despite opposition from some HCW unions and protests across the province, mandates remained effective until July 2024. This study examined the lived experience of HCWs in BC of COVID vaccine mandates, focusing on HCWs’ decision-making process, the mandates’ impact on their lives and livelihoods, and their views on the effects of mandates on patient care.

**Methods:** We performed a reflexive thematic analysis of responses to one open ended question and open-ended entries to closed questions from within a published survey of a convenience sample of HCWs in BC of diverse vaccination status, professions, ages, socioeconomic status, races/ethnicities, and genders. Respondents were recruited through snowball sampling via social media and professional networks of the research team. The study was approved by the York University Office of Research Ethics (No. 2023-389). The article is presented in accordance with the COREQ reporting checklist.

**Results:** Textual data from 90 HCWs were collected from within the initial 166 respondents to the survey. Most respondents were unvaccinated, had been terminated for non-compliance with vaccination mandates, experienced personal losses, and reported negative views on mandates and their impacts on patient care. We identified six themes: 1) policies conflicting with scientific evidence and professional practice; 2) conflicts with medical ethics; 3) unacknowledged or dismissed personal hardships; 4) unacknowledged or dismissed physical harms; 5) discrimination against unvaccinated HCWs and patients; and 6) negative impacts on patient care.

**Conclusions:** This study uncovered a concerning pattern within British Columbia’s healthcare system, with mandatory vaccination leading to substantial social and economic harms for HCWs of all vaccination statuses as well as unvaccinated patients. Mandates appear to have exacerbated labour shortages in the healthcare sector, negatively impacted workplace morale and the quality of patient care, and eroded informed consent. In light of these findings and of available scientific evidence then and now, we conclude that the policy of mandatory vaccination for HCWs has no scientific basis and violates fundamental ethical principles in healthcare practice and policy.

## 1. Introduction

On September 1, 2021, thousands of people from across the Canadian province of British Columbia, including approximately 5,000 in Vancouver, protested COVID-19 vaccine passports and mandatory vaccinations for healthcare workers (HCWs), gathering outside hospitals and city halls. The backlash to, and controversy surrounding, vaccine mandates was not unique to the province - jurisdictions across North America witnessed similar protests over vaccine mandates for HCWs, protests that drew support from individuals working in the healthcare sector, as well as from members of the public who were concerned about expanding vaccine requirements and surveillance (Bains 2021; Shepert 2021; Stahl 2021). At the time of the protests, only one vaccine mandate had been introduced, for long term care and assisted living settings (BC Ministry of Health 2021; Office of the Provincial Health Officer 2022b), yet shortly after, on September 12, another order would extend this requirement to HCWs across all hospitals, mental health facilities, community care, and administrative roles under the provincial and regional health authorities, with a deadline set for October 26, 2021 for HCWs to be vaccinated as a condition of employment in the health sector (Office of the Provincial Health Officer 2022a).

The protests in British Columbia and other provinces in Canada were organized by the Canadian Frontline Nurses, a group that formed upon the World Health Declaration of COVID-19 as a pandemic, and described in a media report as “founded by two Ontario nurses who have promoted conspiracy theories about COVID-19” (CBC News 2021). The demonstrations sparked widespread condemnation from political leaders and public health officials. For instance, Vancouver’s mayor at the time, Kennedy Stewart, called the protesters “kooks” and “fringe lunatics”, and Premier John Horgan and Health Minister Adrian Dix, condemned some protesters, who had been accused of engaging in verbal abuse and physical assaults against compliant HCWs (Lindsay 2021). For his part, the President and CEO of the Canadian Nurses Association (CNA) wrote in a public letter that the actions and views of the Canadian Frontline Nurses did not represent the values of nursing, which he denounced as “anti-science”:

The reckless views of a handful of *discredited* people who *identify as nurses* have aligned in some cases with angry crowds who are putting public health and safety at risk. They have drawn in *anti-science, anti-mask, anti-vaccine*, anti-public health followers whose beliefs align with theirs. For some reason they would have us believe that millions of the best educated health scientists, public health experts, physicians and nurses globally have all missed something they have not. Their outlandish assertions about science would be laughable were they not so dangerous (Canadian Nurses Association 2021) (emphasis added).

The letter went on to say that “anti-public health disinformation threatens to confuse a tired and bewildered public by misrepresenting personal ideology as facts, and science as conspiracy” and finished by reiterating that the Canadian Frontline Nurses “represent everything that we don’t.” These conflicts underscore the polarizing nature of COVID-19 policy, especially vaccine mandates for HCWs in the province. They also illustrate the position of government and public health representatives vis-à-vis those who opposed the policy, whom they framed as a *fringe* and *discredited*, *minority* viewpoint, *identifying as,* rather than real, health professionals - and in opposition to the views of *pro-science, pro-mandate*, health / medical experts and professionals.

Significantly, the comments of government and public health officials also reinforced media accounts at the time – in Canada and beyond - that a *minority* of unvaccinated HCWs as well as members of the public more broadly were responsible for the ongoing COVID-19 crisis (Anthes and Petri 2021; Germany bans unvaccinated people from shops and bars 2021; Pollard and Angus 2021; The Editorial Board 2021). For example, when the vaccine mandate for long term care and assisted living facilities was implemented in August 12, 2021 - about two months before the one including all health facilities - Dr. Bonnie Henry claimed with a sense of urgency that: “We now have eight outbreaks *introduced by unvaccinated people*…and we’ve seen spread both to residents and staff, causing illness but also disruption to the lives of people in long-term care” (Weichel and Daflos 2021)(emphasis added).

Ultimately, vaccine mandates led to an estimated 2,500 HCWs in the province being terminated for non-compliance – over half of them from Interior and Northern Health regions, where labour shortages have resulted in ongoing emergency room closures (DeRosa 2023b); other sources have reported higher numbers - over 4,000 HCWs placed on unpaid leave, with hospitals compelled to cut services due to staff shortages (Government of Canada 2025). The orders remained active until July 26, 2024, when BC’s Provincial Health Officer officially declared the end of the public health emergency and related emergency powers (BC Ministry of Health 2024b). Notably, this was over a year after the WHO – in May 2023 - declared that COVID-19 was no longer a “global health emergency”. As the last jurisdiction in North America to maintain vaccine mandates for HCWs, British Columbia replaced them with a new requirement for them to report their immunization or immune status for COVID-19, influenza and other “critical vaccine preventable diseases” (BC Ministry of Health 2024a).

In truth, by the time mandates were implemented most HCWs in BC had been vaccinated – with, as BC Health Minister Adrian Dix had proffered, “99 per cent of full-time health care workers […] vaccinated for Covid-19, so the number unable to work because they’re unvaccinated” was, according to the Minister, “relatively small” (DeRosa 2023b). It is also true that only a minority of HCWs *openly* challenged the policy. However, the proportion of vaccinated HCWs who may have chosen otherwise had they not been threatened with job termination remains unknown. Equally underexplored is HCWs’ experience of the policy *in their own terms*, meaning free from presuppositions about the policy’s perceived need.

To help fill this gap, we assessed the nature and grounds of vaccination decisions and views on the policy of mandated vaccination among HCWs in British Columbia, by qualitatively examining textual data from one open-ended question and open-ended options within closed questions in a cross-sectional survey exploring these decisions and views. Our study is part of a broader project appraising the impact of the COVID-19 policy response on HCWs and on health systems (Open Science Frame Registration https://osf.io/z5tkp), in turn part of a larger project examining geopolitics, medicalization, and social control in the COVID-19 era (Open Science Frame Registration https://osf.io/84kbr/). We present this article in accordance with the COREQ reporting checklist.

## 2. Methods

We conducted a qualitative reflexive thematic analysis of responses to a single open-ended question (“Are there any other issues that you believe would help us to further understand the impact of the policies on health workers and patient care? If so, please elaborate”) and to open-ended options within closed questions, representing a subset of 90 respondents to a survey of a convenience sample of 166 HCWs in British Columbia. The research team consisted of three female investigators with a joint experience in medical sociology and health services research of over four decades: the lead author is a nonpracticing medical doctor with a Doctorate in Sociology / Notation in Philosophy and employed as a professor of health policy, the first co-author is a health researcher with a Master of Arts in Anthropology, and the second co-author is a health researcher pursuing a Master of Arts in the Health Sciences.

Survey participants were eligible if they were working or had been employed in healthcare before being dismissed due to vaccine mandates. All HCWs in British Columbia, regardless of vaccination status, profession, age, gender, experience, socioeconomic status or race/ ethnicity, were invited to participate. The study was promoted via social media and networks of the principal investigator using a snowball sampling approach to encourage participants to share the invitation with other eligible HCWs. Invitations were redistributed at seven (7) day intervals over the months of May and June of 2024. Details and quantitative results of the survey are reported elsewhere (Chaufan, Hemsing, and Moncrieffe 2025a).

Our analysis was guided by themes identified in a prior qualitative analysis of HCWs’ experience of vaccination mandates in the province of Ontario (Chaufan, Hemsing, and Moncrieffe 2025b) and proceeded in an inductive / deductive fashion – searching for disconfirmation / confirmation of formerly identified themes, while attempting to identify new themes. Data from open-ended responses were extracted from the survey (originally in Google Forms and later transferred to a Word document) and analysis was assisted by Dedoose software.

All authors read the qualitative entries in their entirety, two authors coded them (NH and RM), and the research team met regularly to discuss and interpret findings. The study was approved by the York University Office of Research Ethics (No. 2023-389).

## 3. Results

The textual material represented 90 respondents to the original survey - 78 who replied to the open-ended question and an additional 12 who only elaborated on open-ended options to closed questions. Most respondents reported being unvaccinated or not fully compliant with COVID-19 vaccination requirements. Those who experienced employment termination as a result of vaccine mandates described personal hardships and social and economic losses. Irrespective of vaccination status or current employment, participants overwhelmingly expressed negative views toward vaccination mandates for HCWs. Respondents recounted numerous adverse effects of these mandates, including detrimental impacts on themselves, their colleagues, and their patients. Many conveyed frustrations with policies they perceived as inconsistent with their medical education and practice, institutional protocols, and foundational bioethical principles.

As with the Ontario case, we identified six key themes: 1) policies at odds with scientific evidence, medical training, and usual professional practice; 2) policies at odds with medical ethics; 3) unaccounted or dismissed personal hardships; 4) unaccounted or dismissed physical harms; 5) discrimination based on vaccination status; and 6) negative impacts on patient care. The subsequent section provides an in-depth elaboration and interpretation of each theme to capture respondents’ perspectives, supported by selected verbatim excerpts slightly edited for clarity and coherence.

### 1) Policies at odds with scientific evidence, medical training, and usual professional practice

Respondents described the policy response to COVID-19 as conflicting with their medical training and usual professional practice. Many also reported that they believed that vaccine mandates were unsupported by scientific evidence – particularly the lack of evidence for the ability of vaccination to prevent disease transmission, as well as lack of long-term safety data. One respondent commented that the “data on transmission post vaccination is limited and does not fulfil the logic behind the mandates, nor does the current situational data regarding [COVID-19] in the community validate “emergency orders” (unvaccinated HCW).

Respondents also expressed frustration that mandates continued to treat two doses as adequate for maintaining employment, despite evidence, including from vaccine manufacturers, that two doses did not provide sufficient protection against infection past a few months. For example, one respondent explained that the health authority that employed them required “two doses of the vaccine for a variant that is long gone” and, based on what they concluded was a lack of scientific evidence, argued that “this has nothing to do with our health” (unvaccinated HCW).

Another respondent referred to evidence they had identified, of higher cases of COVID-19, and more missed days of work, among vaccinated and boosted HCWs as compared to unvaccinated HCWs (unvaccinated HCW). Another respondent questioned the scientific rationale for the policy on grounds that remote workers were still mandated to be vaccinated, even if they were not providing “any patient care […], did not come in contact with patients or other employees who worked with patients [and] worked remotely from home, in a different town and different province than where my office was, [yet] was still required to be vaccinated” (partially vaccinated HCW). Yet another respondent, whose comments form the basis of the title of this article, compellingly stated:

2.5 years later I still don’t have my job back. I worked from home. Apparently, my union is still in arbitration. I think it’s a total joke. If all of this was about HEALTH, I’d still be working (unvaccinated HCW).

Respondents also lamented what they perceived to be the lack of evidence informed policy regarding vaccine safety. For example, one respondent stated, “There are no long-term studies, and pregnant women were told to get vaccinated with this new vaccine; did everyone forget about thalidomide or [diethylstilbestrol]” (unvaccinated HCW). Another one explained that they were provided a question-and-answer document which indicated the lack of long-term safety data and noted that vaccination was still required to maintain employment (unvaccinated HCW).

Respondents also criticized how natural immunity acquired from previous infection was ignored in vaccination policies. For example, one respondent noted that “BC does not believe in natural immunity. There is no other option to work as a healthcare professional - you must have two Covid-19 vaccines” (unvaccinated HCW). Another respondent described the “denial of naturally acquired immunity” as a “concerning departure from [evidence informed] policy” (unvaccinated HCW). Yet another described how they sought an exemption from the vaccine mandate based on natural immunity, but ultimately were denied:

I sought exemption from mRNA vaccines as I had already obtained superior natural immunity. I agreed to protein based Novavax that I determined [still] unnecessary but safer platform. I was granted an unpaid 6 month [leave of absence] prior to mandate by employer. Despite this when public health denied my exemption request, I was then given 2 weeks to receive mRNA vaccine or be terminated (unvaccinated HCW).

Many respondents commented that they did not request an exemption even if they wished they had one because they expected to be denied. Finally, respondents called attention to the inconsistency of maintaining a vaccine mandate for HCWs in the province of BC after other provinces, such as Alberta and Ontario, had already rescinded theirs as no longer necessary. In the words of one respondent: “BC is still the only province holding this mandate. What makes them different than Alberta or across Canada where there is no mandate” (unvaccinated HCW).

### 2) Policies at odds with medical ethics

A common theme expressed by respondents was the violation of core bioethical principles, particularly informed consent, which they viewed as coercive measures that compromised individual autonomy and the right to make voluntary health decisions. Notably, respondents concerned about vaccine safety reported that they were assured by employers that the COVID-19 vaccines were safe and effective, even when no evidence to back those claims was ever provided to them. For example, one respondent asserted that “’safe and effective’ [mere claims] are not enough to make an informed decision” (unvaccinated HCW). They also reported that their questions or concerns regarding vaccines were ignored or dismissed (unvaccinated HCW).

Another respondent asked, “What happened to informed consent and bodily autonomy?” (unvaccinated HCW). Yet another one reported having witnessed that patients who were hospitalized for COVID-19 infection were often administered vaccines without consent (unvaccinated HCW). Respondents also noted the loss of patient privacy upon the introduction of mandates, with hospital visitors required to disclose their vaccination status to gain entry:

Patient privacy and confidentiality was thrown out the window as BC implemented a vaccine pass to enter the hospital. The person at the entrance of the hospital would ask for your full name and exactly which department you were going to. No one in the public nor a random person at the door needs to know your business (unvaccinated HCW).

The lack of privacy and confidentiality of personal medical data also extended to HCWs. One respondent explained “Although, annually you needed to manually report your flu vaccination status, with COVID-19 the health authority just went into your personal records and looked it up. No confidentiality for healthcare workers” (unvaccinated HCW).

### 3) Unacknowledged or dismissed personal hardships

Another salient theme was the lack of recognition or outright disregard, by the authorities imposing the mandates, of the personal hardships and struggles faced by HCWs. Respondents described being terminated for non-compliance with mandatory vaccination, often without any acknowledgement of how deeply such decisions impacted their lives, and often after years of dedicated service. Many identified a wide range of personal and economic setbacks due to the mandates and shared stories of severe financial strain that followed their implementation. These included losing access to severance or employment insurance, depleting lifelong savings, and being dependent on financial help from family members or friends to survive. As well, many respondents reported being “forced into retirement” due to the pressure of vaccination or losing employment, with one respondent noting that the “number of those that retired early to avoid vaccination-or-termination is not reported or understood” and that “current gaps in healthcare are bigger based on those unreported numbers” (unvaccinated HCW).

One respondent said that the policy of mandated vaccination had been an “absolutely indescribable disaster affecting every area of one’s life” (unvaccinated HCW). Examples of these life-altering circumstances were provided by another respondent, who explained: “People I know sold homes, moved to different provinces or even to the United States - making major life alterations because of government policy” (unvaccinated HCW). Another one described their experience of being placed on unpaid leave, losing access to health benefits, and needing to take on contract work to earn an income:

My ex-husband was dying during all of this, and I had to leave my position as an addictions counsellor or risk termination for not being vaxxed. I have been on unpaid leave since October 2021 and continue to do contract work in order to have an income. My contract work used to be my side gig. Now it is my income. I have to pay for my own benefits in order to maintain even dental for myself (unvaccinated HCW).

Many respondents also reported being terminated *with cause*, meaning for reasons considered legally valid, and for which the employee is made responsible - therefore deserving to be denied employment insurance benefits. One respondent explained their experience of this dilemma:

“There were no layoffs, no access to [Employment Insurance]; it was immediate termination with cause. I was breastfeeding my toddler” (unvaccinated HCW). Others reported being no longer interested in remaining in the health sector out of a sense of distrust, loss of respect, and personal hurt. For instance, one respondent reported having experienced “moderate depression and anxiety” over losing their nursing career” yet also reported that they still would not want to return to their previous role “due to the prejudice [I experienced] by colleagues for not being vaccinated” (unvaccinated HCW). Another one reported their experience and observation of “loss of income resulting in loss of homes, separation of marriages, distrust in the healthcare system” (unvaccinated HCW). Yet another one compellingly described how mandates had left a “permanent scar”:

What “public health” did during the pandemic changed me as a person for life. I have managed to find a new way, but this has left [in me] a permanent scar. I now don’t trust our health system; I now see and understand the relationship pharmaceutical companies have with our medical system (fully vaccinated HCW).

Similarly, another respondent reported that while at the time of the survey they were doing better than when mandates were in effect, they still “grieved” their job and remained “emotionally distraught” over how they had been treated at the height of the mandated vaccination period (unvaccinated HCW).

### 4) Unacknowledged or dismissed physical harms

Another prominent theme was the strong belief held by many respondents that the increase in illness and disability among vaccinated patients – which they felt were often ignored, dismissed, or denied - were linked to the COVID-19 vaccine. For example, one respondent pointed out their observation about the silence regarding adverse events among patients and HCWs:

People are being diagnosed with myocarditis and pericarditis and various cancers and there is no mention of the vaccines probably causing this. This deceit is very stressful to health workers and very damaging to patients, who no longer know who to trust in health care (unvaccinated HCW).

Respondents also observed health issues among patients that they believed were linked to the COVID-19 vaccine and expressed frustration that these concerns were either not investigated or quickly dismissed by most colleagues as unrelated to vaccination. For example, one participant reported both directly caring for, and knowing of, patients who were vaccine injured, and yet were pressured into receiving additional vaccine doses:

I worked with rehab patients who were injured as a result of the shot, and it was documented, yet I was muzzled to give them information that would help them get better and the unit continued to coerce patients to get more shots despite their injury. The rehab docs and [general practitioners] did not consider some injuries to be due to the shot or were not allowed to even consider [the vaccine to be the cause] if they thought so. One doctor had a vaxx injury himself but could not give a recommendation to a patient on not taking another shot even though she herself had sustained a vaxx injury (unvaccinated HCW)

Similarly, another HCW reported witnessing adverse effects among patients following vaccination:

I saw our patients being adversely affected when they rolled out the vaccine and encouraged nurses to document the adverse effects. We witnessed stable patients fail and die rapidly post-vaccine, but no one did anything about it (partially vaccinated HCW).

In addition, some vaccinated respondents experienced adverse effects post COVID-19 vaccination, including one respondent who was refused an exemption despite experiencing an adverse reaction post vaccination (partially vaccinated HCW). Another respondent described their severe adverse reactions following each dose of the COVID-19 vaccine, as follows:

After the first dose, I had severe nosebleeds and blood clots passing through my nose for about two weeks. I thought I was going to have a stroke and die. So, to me, it was a life-threatening reaction at the time! I do have an auto immune disease (partially vaccinated HCW).

### 5) Discrimination according to vaccination status

Many respondents vividly described the discrimination they experienced, from employers, the larger society, and even colleagues, due to their vaccination status. Some respondents commented that they were fired for non-compliance with vaccine mandates, even though they had worked with patients, including those infected with COVID-19, through 2020, before vaccines were available, and into 2021, before they were mandated. For example, one respondent described what they saw as the irrationality of vaccine mandates for HCWs even when they had been repeatedly exposed to the virus working in the pre-vaccine and pre-mandate era:

“You work through the first two years of a pandemic with limited knowledge of the virus and continually see patients in person, and then once the vaccine mandates come out and you don’t take the vaccine you’re a monster and an uneducated healthcare worker” (unvaccinated HCW).

Many respondents felt that the discrimination they experienced or observed was too much to bear, causing them to not only lose trust in the system but also to leave their profession. For example, one respondent reported that they would not return to work in healthcare because of the “prejudice” they experienced from colleagues (unvaccinated HCW). Another respondent explained that: **“**the health care system has lost many highly skilled professionals due to them being terminated…and has degraded significantly as a result; coercive policies have made the industry undesirable to participate in” (unvaccinated HCW).

A related theme was the increased hostility and loss of camaraderie within the profession because of vaccine mandates. One respondent shared: “These policies really divided everyone in a very negative way. HCWs often need to work as team members; [mandates] did not help anyone” (unvaccinated HCW). Another respondent reported how vaccine mandates contributed to a toxic and discriminatory environment for both patients and HCWs:

The divide it created in the hospital was sickening - witnessing nurses and doctors and other medical staff make fun of unvaccinated patients and saying things like they don’t deserve to be seen before anyone else was sickening. To be part of the ostracization and witness it felt like the holocaust. One of our RN’s went on early mat leave because she was constantly being harassed by one of the doctors to get vaccinated while being pregnant. This doctor also made it known she did not want to see unvaccinated patients (unvaccinated HCW).

One seemingly aggrieved respondent observed that “Dr. Henry’s constant use of the phrase “an unvaccinated HCW represents a public health hazard” and the “language used to portray the unvaccinated as selfish, ignorant and backward created significant animosity” within the profession and among the public towards unvaccinated persons more broadly (unvaccinated HCW). Many respondents also observed serious discrimination against patients who were unvaccinated, discrimination that in their view significantly impacted the quality of patient care. For example, one HCW described some colleagues behaviour as “completely unprofessional”, noting that “many nurses should lose their license for how they talked about their patients’ choice to not vaccinate” (unvaccinated HCW). Another one observed that the “discrimination against the unvaccinated was extreme, with many clinics turning away patients, even as they received “young adults who were suicidal because the route to education in their desired health profession closed unless they were vaccinated” (unvaccinated HCW).

Finally, respondents also chronicled how unvaccinated people were denied access to visit loved ones in hospital care, negatively impacting the quality of care of patients of *all* vaccination statuses, as well as other practice changes that they considered unnecessary and inhumane, such as mandating masks during delivery, or banning unvaccinated partners from being present at the moment of delivery:

Humanity was lost. Women were told to wear a mask while delivering their babies; spouses / partners were not allowed in for deliveries if they did not have a vaccine pass (unvaccinated HCW).

### 6) Negative impacts on patient care

In addition to observing physical harms and serious instances of discrimination suffered by patients, respondents also reported that vaccine mandates exacerbated staffing shortages, as HCWs were either placed on extended unpaid leave or dismissed, ultimately degrading the quality of patient care. For example, one respondent was aware of “a few local nursing friends” who would soon lose their licenses because they were unable to meet their hours requirement “because they are unemployable due to their unvaccinated status” (unvaccinated HCW). The same respondent also described the case of HCWs who were barred from working as caring and eager to work, noting that “some of these nurses had another 20+ years left in them to work, and loved their jobs and those they cared for” (unvaccinated HCW).

Another respondent expressed shock that the Ministry of Health had “completely failed to assess the risks posed by understaffing and service closures because of the mandates” (unvaccinated HCW). Yet another one remarked that “the impact trickles down to ordinary citizens” because “the fewer healthcare workers, the fewer staff will be available to address the healthcare needs of patients” (unvaccinated HCW). Finally, one participant wrote that: “healthcare in BC is poisoned” with many HCWs resenting their employers “for being coerced” to be vaccinated against their will, which they believed “can only reflect on the care they provide” (unvaccinated HCW).

Importantly, many respondents described how healthcare priorities had shifted entirely towards COVID-19 to the neglect or dismissal of other health priorities. For example, one respondent, a radiographer, described how COVID-19 policy changes caused patients who were requiring imaging to use portable machines, which exposed them to more radiation and unnecessary risks, stating that “it felt like EVERYTHING was about COVID, when it didn’t need to be […]; it burned our staff our very quickly and degraded patient care” (unvaccinated HCW). Another respondent also observed the excessive attention to COVID-19 to the detriment of multiple pressing health issues in the province, such as substance use and addictions, either prior to, exacerbated by, or resulting from, the COVID policy response itself:

The spike in overdose deaths in Vancouver [most populated BC city] directly relates to the service restrictions initiated by covid protocols prioritizing one group over all others. I am certain more overdose deaths have happened than covid deaths. We were told pre-covid there was no money for anything to improve care for substance users - then boom, covid happens and suddenly we have all the money in the world, [but only] for covid and nothing else. Hypocrisy (fully vaccinated HCW).

## 4. Discussion

All respondents in this study completing the open-ended question or filling out open ended options in closed questions opposed vaccine mandates - with many describing emotional, social and financial hardships following the implementation of the policy. Most respondents chose to not comply, and as a result experienced job loss, financial strain, conflict with colleagues, friends and family, and mental and emotional distress. Respondents who were terminated felt alienated from their profession and institutions they once trusted and served. They also criticized the mandates for disregarding conflicting scientific data, particularly regarding viral transmission, vaccine safety, and natural acquired immunity, and felt that their clinical expertise and safety concerns had been routinely dismissed in favour of a narrative that promoted the vaccine as universally “safe and effective.” Additionally, many respondents observed how such policies compromised both accessibility and quality of care, fostering discrimination against unvaccinated HCWs and patients. Finally, many reported that mandates violated the core ethical principle in medicine of informed consent. All these findings echoed those in a similar survey conducted with HCWs in the province of Ontario, where vaccination mandates were implemented across healthcare settings (Chaufan, Hemsing, and Moncrieffe 2025c).

In alignment with the current qualitative findings, our quantitative analysis of the data that this article draws from revealed that of 166 survey respondents of mixed vaccination status, a large majority (81%) experienced anxiety or depression due to mandated vaccination, and close to one fourth (23.5%) reported suicidal thoughts (Chaufan, Hemsing, and Moncrieffe 2025a). As well, a large majority (73%) experienced a decrease of their income due to vaccination policies (a combination of layoffs, resignations, and early retirements), and most (84%) reported that their personal relationships had suffered, about one third (34%) reported negative impacts on their physical health, and over half (58%) experienced deterioration of their mental health.

Significantly, most (93%) respondents were offered no accommodations or alternatives to vaccination, whether for medical, conscience, or religious reasons, and most (90%) reported feeling unfree to choose whether or not to be vaccinated. In relation to the potential impact of the policy on patient care via the exacerbation of the staffing crisis in the province, close to half (45%) of respondents were no longer interested in remaining in the industry. In summary, our survey indicates that vaccination policies had an overall negative impact on the physical, emotional and social well-being of HCWs, particularly – albeit not only - those who did not comply with mandates, as well as on health services and concomitantly, patient care.

While the Provincial Health Officer of BC rescinded the COVID-19 public health emergency and the vaccination mandate in healthcare settings, challenges remain: for instance, the provincial government has now introduced a requirement for all HCWs in public healthcare settings to report their vaccination status for COVID-19 and influenza, as well as their immune status – through vaccination or naturally acquired - for measles, mumps, rubella, hepatitis B, whooping cough (pertussis), and chicken pox (varicella) (BC Ministry of Health 2024a). As per BC authorities, the government’s move “requires the immune status of health-care workers to protect both patients and workers [and is] part of a system that can help to prevent outbreaks and manage them when they do happen quickly and effectively” (BC Ministry of Health 2024a).

Furthermore, HCWs in British Columbia have voiced strong concerns about the Health Professions and Occupations Act (HPOA), formerly Bill 36, which was passed in November 2022 and is set to take effect in 2025. The legislation is informed by a 2018 report by independent expert Harry Cayton - who identified governance issues and the need for a complete overhaul of health regulation in the province - which cited as evidence incidents like the spread of an “anti-vaccine video” by a BC College of Chiropractors board member (Cayton 2018). The Health Professions and Occupations Act aims to “modernize” regulation by consolidating 15 health colleges into six and shifting board appointments to government appointees, half of whom must be non-licensed public members (BC Government News 2022).

For his part, BC Health Minister has asserted that the bill was informed through one of the “most extensive consultation processes in the government’s history” (DeRosa 2023a) and that it will “streamline the process to regulate new health professions, provide stronger oversight [and] protect patient care” (BC Government News 2022). Critics, however, have warned that the bill is a top-down measure implemented undemocratically and likely to further undermine patient care - only 233 of the 645 sections were discussed, and many HCWs, including the organization Doctors of British Columbia, have warned that it will exacerbate recruitment challenges amid ongoing labour shortages (Hsiang 2024). In the words of one physician, “nurses are […] actually leaving our community to go elsewhere, and physicians, same thing….We need to make the grounds amenable to healthcare workers so they will want to stay” (Kingston 2024). Another critic, a HCW, has argued that the Health Professions and Occupations Act fosters mistrust, noting that it creates an environment that will not “attract doctors or other healthcare professionals to B.C.” (Lush 2023).

So far, there has been a petition, and calls, for the repeal or delay of the Health Professions and Occupations Act, though the government insists that the changes it will implement are essential for improving patient safety and restoring public confidence in healthcare (Oral Health 2025). However, the new reporting and regulatory requirements may be replicating the policy tensions and HCWs’ experiences observed with COVID-19 vaccination mandates - top-down countermeasures that ignore significant pockets of resistance among HCWs, greater distrust in public health authorities among sectors of the public, erosion of informed consent and confidentiality between healthcare professionals and patients, and a greater number of HCWs discouraged from maintaining or taking up practices in the province (BC College of Family Physicians 2023; DeRosa 2023a; Hsiang 2024; Kingston 2024; Lush 2023; Oral Health 2025). Thus, while the orders restricting unvaccinated HCWs from employment have been rescinded, the health surveillance infrastructure appears to be expanding, with likely negative effects on the ongoing shortage of physicians and nurses (BC Nurses Union 2023; Judd and Stanton 2025; Li et al. 2023).

Meanwhile, HCWs in British Columbia have continued to challenge COVID vaccine mandates, largely on ethical grounds – infringement of constitutional rights, and freedoms of conscience, religion, and personal security. However, to date, courts have largely aligned with the position of government and public health officials, justifying mandated vaccination for HCWs based on the alleged scientific evidence available at the time (Hoogerbrug v. British Columbia 2024; Notice of Civil Claim: Jedediah Ferguson and Terri Perepolkin vs. Dr Bonnie Henry 2023). Still, the courts have requested Provincial Health Officer of BC, Dr. Bonnie Henry, to reconsider the order for HCWs who worked remotely and did not provide patient care (Hoogerbrug v. British Columbia 2024). In response, on August 28, 2024, Dr. Henry issued a reconsideration decision, confirming her position of not approving exemptions for remote and administrative workers based on three reasons: 1) during a public health emergency, remote workers could be required to be physically present including through formal deployment; 2) COVID-19 vaccination “was and remains” effective in preventing severe illness and is needed to protect the healthcare system; and 3) having an exception in the policy for remote workers would have been impractical at a time given limited public health resources (Office of the Provincial Health Officer 2024).

At this point we should note the problematic assumptions underlying government claims, which omit important scientific counter evidence: for example it was well determined early on that COVID-19 vaccines were unable to stop viral transmission (Shrestha et al. 2023; Singanayagam et al. 2021), including among fully vaccinated people in healthcare settings (Mateos-Nozal et al. 2021; Park et al. 2022; Public Health Agency of Canada 2022). This was, and continues to be, the fundamental scientific rationale underlying mandate implementation. Mandate policies have also dismissed accumulating scientific evidence of adverse events – from mild to life-threatening to lethal – also available early in the COVID vaccination campaign (Bareiß et al. 2022; Faksova et al. 2024; Farah et al. 2023; Filippatos et al. 2021; Ponticelli et al. 2021; Seneff et al. 2022; Yamamoto 2022). Importantly, and as reported by respondents in this study, mandated vaccination infringes on the right to informed consent - established in seminal documents (Shuster 1997; UNESCO 2005; World Medical Association 1964) – which includes the right to be fully informed about the risks, benefits, and alternatives to any medical intervention, including the alternative to refuse treatment free from any form of coercion or threat.

This study has limitations inherent to the exploratory nature of the research project and of qualitative research more broadly. Although HCWs of any vaccination status were invited, more unvaccinated than vaccinated HCWs chose to participate. Therefore, there is no evidence that the results reflect the experiences of all or even most HCWs in the province, most of whom were vaccinated. One way to interpret the observation of high vaccine uptake among HCWs in the province is that most of them supported mandates. On the other hand, this same observation could be interpreted as meaning that while only a minority of HCWs *openly* challenged the policy, a significant number accepted to get vaccinated under coercion, to avoid a punishment that few of them could afford - unpaid leave, and eventually termination, often with no right to compensation or unemployment benefits, as reported by several respondents in our study.

While either fully free consent or coercion could be the correct explanation to the observed high vaccination uptake among HCWs, the coercive nature of the policy is suggested by independent evidence: one national, cross-sectional survey of 5,372 HCWs conducted by the Canadian polling company Ipsos for the Public Health Agency of Canada - to our knowledge the largest and methodologically strongest of its kind in the country – found that vaccine mandates were a major reason for vaccination, with 53% of healthcare professionals, 46% of allied health workers, and 47% of auxiliary health workers citing the threat of job loss as a key factor (Ipsos & Public Health Agency of Canada (PHAC) 2023). Similarly an Italian study found that most HCWs surveyed shifted their stance from rejecting to “accepting” vaccination due to the threat of work restrictions if they did not (Costantino et al. 2022).

Our methodological limitations notwithstanding, our research fills an important gap, as most existing studies on the impact of vaccination mandates have been conducted with primarily vaccinated HCWs, thus excluding by design the voices of dissidents. For example, a systematic review investigating HCWs’ attitudes toward mandatory vaccination reported the potential for bias due to high vaccination rates among participants in the included studies (Politis et al., 2023). In contrast to most studies, we did not recruit within medical establishments, which would have led us to reach only vaccinated HCWs – the only ones who remained employed – thus the limitations of our recruitment methods may have turned out to be a strength, as social media allowed us to reach unvaccinated HCWs, most negatively affected by vaccination policies and for this reason eager to have their experiences and views considered.

## 5. Conclusions

In conclusion, our findings indicate that COVID-19 vaccination mandates contributed to a healthcare system in British Columbia, Canada, that discriminated against HCWs who did not receive the two doses required to be considered “fully vaccinated” to retain employment (Wong 2022). As discussed in the introduction, proponents of vaccination mandates have argued that the differential treatment of unvaccinated HCWs was justified and necessary to protect patients, other HCWs, and even dissidents themselves, and legal attempts to challenge mandates have been largely unsuccessful on similar premises. However, we have reported substantial scientific evidence challenging these assertions. We have also reported on the substantial social, emotional, and economic harms inflicted by a policy that appears to have exacerbated critical issues within the healthcare sector, including labour shortages, the deterioration of workplace morale, a decline in the quality of patient care, and the erosion of informed consent. In light of these findings and of our analysis of publicly available medical literature, we conclude that the practice of mandatory vaccination within healthcare settings has no scientific basis and violates fundamental ethical principles in healthcare practice and policy.

## Data Availability

Data are not available due to confidentiality agreement with survey respondents. Summarized, quantitative data are available upon reasonable request to the authors.

## Acknowledgments

CC thanks the many professional and lay organizations, students, trainees, and friends who have afforded spaces of reflection and debate over the past years, and especially Julian Field, for his editorial feedback and support. NH thanks her family and friends for their encouragement and support, and Dr. Chaufan for her mentorship. RM thanks her friends, family, and Dr. Chaufan for their support and guidance. All authors are grateful to the participants for sharing with us their life experiences and making this study possible.

## Author Contributions (CRediT taxonomy)

Conceptualization: Claudia Chaufan

Methodology: Claudia Chaufan and Natalie Hemsing

Writing – Original Draft: Claudia Chaufan

Writing – Review & Editing: Claudia Chaufan, Natalie Hemsing, Rachael Moncrieffe

Investigation: Natalie Hemsing and Claudia Chaufan

Formal Analysis: Claudia Chaufan, Natalie Hemsing & Rachael Moncrieffe

Project Administration: All authors

## Reporting Checklist

The authors have completed the COREQ reporting checklist.

## Ethical Statement

The authors are accountable for all aspects of the work in ensuring that questions related to the accuracy or integrity of any part of the work are appropriately investigated and resolved. The study was approved by the York University Office of Research Ethics (No. 2023-389). Potential study participants were provided an information letter and consent form, including details on the study aims, methods, potential benefits and risks, and information about confidentiality and consent. They were informed of their right to withdraw consent at any time without consequences. The online survey questions were only accessible to participants after providing their freely informed consent.

