## Supplementary material for "“If all of this was about HEALTH, I’d still be working”: Lived Experiences of Covid-19 Vaccine Mandates of Healthcare Workers in British Columbia, Canada": NA

### Consolidated criteria for reporting qualitative studies (COREQ): 32-item checklist

| No | Item | Guide questions/description | Reported on Page Number/Line Number | Reported on Section/Paragraph |
|---|---|---|---|---|
| **Domain 1: Research team and reflexivity** | | | | |
| Personal Characteristics | | | | |
| 1 | Interviewer/facilitator | Which author/s conducted the interview or focus group? | p.8 | Methods |
| 2 | Credentials | What were the researcher's credentials? e.g. *PhD, MD* | p.8 | Methods |
| 3 | Occupation | What was their occupation at the time of the study? | p.8 | Methods |
| 4 | Gender | Was the researcher male or female? | p.8 | Methods |
| 5 | Experience and training | What experience or training did the researcher have? | p.8 | Methods |
| Relationship with participants | | | | |
| 6 | Relationship established | Was a relationship established prior to study commencement? | N/A | N/A |
| 7 | Participant knowledge of the interviewer | What did the participants know about the researcher? e.g. *personal goals, reasons for doing the research* | p.8 | Methods |
| 8 | Interviewer characteristics | What characteristics were reported about the interviewer/facilitator? e.g. *Bias, assumptions, reasons and interests in the research topic* | p.8 | Methods |
| **Domain 2: study design** | | | | |
| Theoretical framework | | | | |
| 9 | Methodological orientation and Theory | What methodological orientation was stated to underpin the study? e.g. *grounded theory, discourse analysis, ethnography, phenomenology, content analysis discourse analysis, ethnography, phenomenology, content analysis* | p.7 | Methods |
| Participant selection | | | | |
| 10 | Sampling | How were participants selected? e.g. *purposive, convenience, consecutive, snowball* | p.8 | Methods |
| 11 | Method of approach | How were participants approached? e.g. *face-to-face, telephone, mail, email* | p.8 | Methods |
| 12 | Sample size | How many participants were in the study? | p.8 | Results |
| 13 | Non-participation | How many people refused to participate or dropped out? Reasons? | p.8 | Results |

| | | | | |
|---|---|---|---|---|
| Setting | | | | |
| 14 | Setting of data collection | Where was the data collected? e.g. *home, clinic, workplace* | p.8 | Methods |
| 15 | Presence of non-participants | Was anyone else present besides the participants and researchers? | Not in person study | Methods |
| 16 | Description of sample | What are the important characteristics of the sample? e.g. *demographic data, date* | p.8 | Methods |
| Data collection | | | | |
| 17 | Interview guide | Were questions, prompts, guides provided by the authors? Was it pilot tested? | p.7 | Methods |
| 18 | Repeat interviews | Were repeat interviews carried out? If yes, how many? | NA not in person study | Methods |
| 19 | Audio/visual recording | Did the research use audio or visual recording to collect the data? | NA not in person study | Methods |
| 20 | Field notes | Were field notes made during and/or after the interview or focus group? | NA not in person study | Methods |
| 21 | Duration | What was the duration of the interviews or focus group? | NA not in person study | Methods |
| 22 | Data saturation | Was data saturation discussed? | NA, based on survey | Methods |
| 23 | Transcripts returned | Were transcripts returned to participants for comment and/or correction? | NA, not in person study | Methods |
| **Domain 3: analysis and findingsz** | | | | |
| Data analysis | | | | |
| 24 | Number of data coders | How many data coders coded the data? | p.8 | Methods |
| 25 | Description of the coding tree | Did authors provide a description of the coding tree? | p.8 | Methods |
| 26 | Derivation of themes | Were themes identified in advance or derived from the data? | p.8 | Methods |
| 27 | Software | What software, if applicable, was used to manage the data? | p.8 | Methods |
| 28 | Participant checking | Did participants provide feedback on the findings? | NA not in person study | Methods |
| Reporting | | | | |
| 29 | Quotations presented | Were participant quotations presented to illustrate the themes/findings? Was each quotation identified? e.g. *participant number* | p.10 - 21 | Results |
| 30 | Data and findings consistent | Was there consistency between the data presented and the findings? | As per quotations | Results |
| 31 | Clarity of major themes | Were major themes clearly presented in the findings? | p.10 - 21 | Results |
| 32 | Clarity of minor themes | Is there a description of diverse cases or discussion of minor themes? | p.10-21 | Results |

**From:** Tong A, Sainsbury P, Craig J. Consolidated criteria for reporting qualitative research (COREQ): a 32-item checklist for interviews and focus groups. Int J Qual Health Care. 2007;19(6):349-57

Please leave this space alone as it will be supplemented by the editorial office when needed.
